# Significance of Wastewater Surveillance in Detecting the Prevalence of SARS-CoV-2 Variants and Other Respiratory Viruses in the Community – A Multi-Site Evaluation

**DOI:** 10.1101/2023.01.05.23284236

**Authors:** Majid Khan, Lin Li, Laura Haak, Shannon Harger Payen, Madeline Carine, Kabita Adhikari, Timsy Uppal, Paul D. Hartley, Hans Vasquez-Gross, Juli Petereit, Subhash C. Verma, Krishna Pagilla

**Affiliations:** School of Medicine, University of Nevada, Reno, NV, 89557, USA; Department of Microbiology and Immunology, University of Nevada, Reno School of Medicine, MS320, Reno NV, 89557, USA; Department of Civil and Environmental Engineering, University of Nevada, MS258, Reno, NV 89557, USA; Nevada Genomics Center, University of Nevada, Reno, NV, 89557, USA; Nevada Bioinformatics Center, University of Nevada, Reno, NV, 89557, USA

**Keywords:** WBE, Diagnostics, RVP, Variants, Coinfection, AMRs

## Abstract

Detection of severe acute respiratory syndrome coronavirus 2 (SARS-CoV-2) viral genome in wastewater has proven to be useful for tracking the trends of virus prevalence within the community. The surveillance also provides precise and early detection of any new and circulating variants, which aids in response to viral outbreaks. Site-specific monitoring of SARS-CoV-2 variants provides valuable information on the prevalence of new or emerging variants in the community. We sequenced the genomic RNA of viruses present in the wastewater samples and analyzed for the prevalence of SARS-CoV-2 variants as well as other respiratory viruses for a period of one year to account for seasonal variations. The samples were collected from the Reno-Sparks metropolitan area on a weekly basis between November 2021 to November 2022. Samples were analyzed to detect the levels of SARS-CoV-2 genomic copies and variants identification. This study confirmed that wastewater monitoring of SARS-CoV-2 variants can be used for community surveillance and early detection of circulating variants and supports wastewater-based epidemiology (WBE) as a complement to clinical respiratory virus testing as a healthcare response effort. Our study showed the persistence of the SARS-CoV-2 virus throughout the year compared to a seasonal presence of other respiratory viruses, implicating SARS-CoV-2’s broad genetic diversity and strength to persist and infect susceptible hosts. Through secondary analysis, we further identified antimicrobial resistance (AMR) genes in the same wastewater samples and found WBE to be a feasible tool for community AMR detection and monitoring.

**HIGHLIGHTS/KEY FINDINGS:**

- WBE better represents regional virus prevalence under declining testing and clinical reporting rates.
- SARS-CoV-2 virus was present throughout the year compared to a seasonal prevalence of other respiratory viral pathogens.
- SARS-CoV-2 variant monitoring via WBE can provide insights into regionally co-circulating respiratory viruses, causing flu-like symptoms.
- WBE-AMR gene monitoring is feasible and provides valuable insight into regional AMRs.
- Our workflow enables the estimation of the relative proportion of different variants in any pooled samples.

**GRAPHICAL ABSTRACT:** 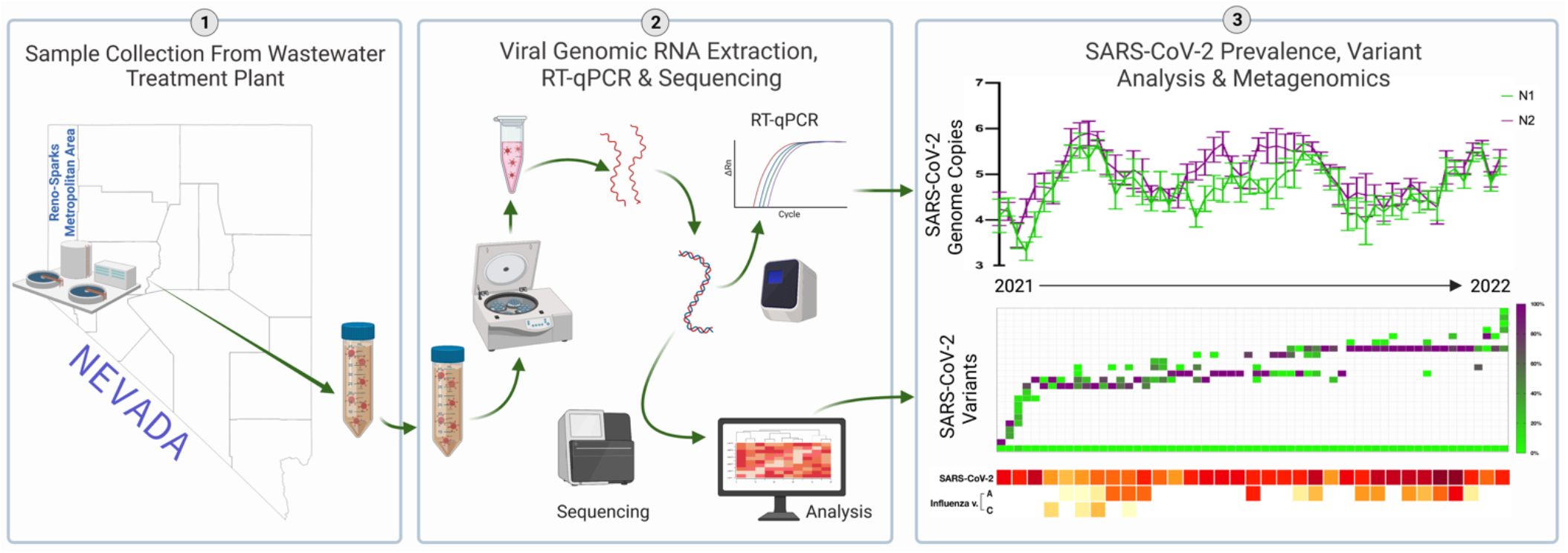

## INTRODUCTION

The Severe Acute Respiratory Syndrome Coronavirus 2 (SARS-CoV-2) virus, responsible for the Coronavirus Disease of the 2019 (COVID-19) pandemic, continues to be prevalent globally. In the United States of America (USA), case numbers are approaching 100 million, with a death rate that has surpassed 1.09 million persons as of 12/05/2022.^1^ Infectious diseases are a significant threat to global and public health, with many drivers causing increased spread and transmission.^2^ Following CDC direction, many clinical and public health laboratories, academic institutions, and private sectors have contributed significant efforts to monitor virus evolution through next-generation sequencing (NGS) technologies, which have supported tracing efforts and assisted in research on transmission dynamics and host response.^3^ However, alternative and complementary technologies are also necessary for this effort as there exist barriers to the accessibility of sequencing technologies (i.e., rising costs and sizeable necessary sequencing volumes).^4,5^ Furthermore, leveraging the SARS-CoV-2 monitoring to include co-occurring pathogens in the community, including other respiratory viruses that cause similar symptoms, is critical to accurately determine community health and disease prevalence.

SARS-CoV-2 viral particles are present in the feces of virus carriers and subsequently shed into sewage systems and downstream water reclamation facilities (WRFs).^6,7^ Thus, wastewater-based epidemiology (WBE) within these sewage networks and WRFs can provide valuable insight into virus prevalence and variant evolution in sewershed.^8,9^ Moreover, WBE shows a low risk of virus infectivity to personnel, although caution should still be used.^10,11^ Wastewater samples allow for the anonymous quantification of SARS-CoV-2 viral RNA from individuals in a defined sewershed, representing the overall spatial and temporal prevalence of SARS-CoV-2 in a community at a given time point.^12-14^ WBE has been shown to provide earlier detection of circulating respiratory viruses and variants of concern (VOCs), which can appear days to weeks before clinical cases are seen in healthcare settings (e.g., detection of B.1.1.519 preceding the Omicron wave).^15-17^ It has also been shown that WBE of SARS-CoV-2 has a strong correlation with clinically diagnosed case numbers;^12^ further, the WBE methodology is not dependent on external factors (e.g., testing and reporting rates, reporting lag, and reporting biases).^12,18,19^

In this study, we captured and enriched SARS-CoV-2 in the wastewater, reported site-specific sequencing and variant analysis, and described the implications of these data. Further, we performed a respiratory viral pathogens (RVP) panel analysis for other respiratory disease-causing viruses in these samples. Use of widespread antimicrobial agents during the pandemic could potentially trigger higher antimicrobial resistance (AMR) occurrence in the community. Thus, to contribute additional insights into community health risks due to other pathogens, this study correlated the presence of the AMR genes in wastewater with SARS-CoV-2 and other respiratory viruses. The goal was that analyzing SARS-CoV-2, other RVPs, and AMRs in the wastewater may allow for increased knowledge and clarity of impending infection waves, which may provide insight into the prevalence and evolution of respiratory viruses in the community and inform public health decision-making.

## MATERIALS AND METHODS

### 1.1 Sample and Study Sites

This study was conducted in the Reno-Sparks metropolitan area in Nevada (NV), USA, between November 2021 and November 2022. Seven local sub-sewersheds and influent (pre-treatment) from three water reclamation facilities (WRF) sites were included in the study, representing wastewater collected from about 0.4 million residents. The WRFs ranged from the southern Truckee Meadows region to the northern Reno-Stead region and included the Truckee Meadows WRF (TMWRF), the South Truckee Meadows WRF (STMWRF), and the Reno-Stead WRF (RSWRF). The Truckee Meadows WRF (TMWRF) represents sewershed catchments from both Reno and Sparks, NV, serving over 205,000 and 115,000 residents, respectively, and has an approximate 121,000 m^3^/day flow rate (∼30 million gallons/day), accounting for about 80% of Washoe County’s wastewater. The South Truckee Meadows WRF (STMWRF) serves over 52,000 residents and has an approximate 96,000 m^3^/day flow rate (∼2.5 million gallons/day). The Reno-Stead WRF (RSWRF) serves over 18,000 residents and has an approximate 64,000 m^3^/day flow rate (∼1.6 million gallons/day). In addition to these WRFs, we also analyzed influent from two hotel-casinos to represent Travel-Influenced Sites and combined influent from three sewer sub-catchments with inflow exclusively from residential housing areas (approximately 500 residential units each) to represent Sub-Neighborhoods (Sparks, NV). Two elementary schools, one in Reno and one in Sparks, NV, were also included to represent Elementary Schools in the region.

### 1.2 SARS-CoV-2 Specimen Collection and Quantification in Wastewater

The methodology for virus enrichment and quantification in wastewater was performed as described in our previous study.^20^ Briefly, 1 liter (L) of untreated wastewater was obtained after preliminary treatment from facilities between 9:00 a.m. to 12:00 noon and transported directly to the laboratory on ice. Samples were kept at 4 °C until further treatment. Samples were centrifuged at 3000×g for 15 min, and the resulting supernatants were sequentially filtered through 1.5, 0.8, and 0.45 μm sterile membrane filters to remove debris and large particles. The resulting supernatant was used to concentrate the viruses. The virus concentration was performed via ultrafiltration using 100 KDa Amicon® Ultra-15 Centrifugal Filter Cartridge Units (Millipore Sigma, St. Louis, MO, USA). Depending on the wastewater virus concentration levels, we processed 60 mL of samples (i.e., to concentrate the viruses to a detectable level). After ultrafiltration, a cartridge of ∼ 500 μL of the concentrate was collected and stored at −80°C for downstream analysis (unless analyzed that day).

The total RNA from the concentrated samples was extracted using an AllPrep PowerViral DNA/RNA kit, following the protocol provided (QIAGEN, Inc., Germantown, MD, USA). Reverse transcription and quantitative polymerase chain reactions (RT-qPCR) were completed using the CFX96 Touch Real-Time PCR Detection System (BioRad, Hercules, CA, USA). N1 and N2 primers and probes were used for the RT-qPCR assay, per US-CDC recommendations.^21^ The RT-qPCR was conducted by SARS-CoV-2 RT-qPCR Kits for wastewater (Promega, Madison, WI, USA) according to the kit instruction manual. Briefly, each reaction contained 10 μL GoTaq® wastewater Probe qPCR MasterMix (x2), 1 μL N1 and N2, respectively, and PMMoV Primer/Probe/IAC Mix (x20), 0.2 μL GoScript® Enzyme Mix (x50), and 5 μL of the total genomic RNA template, into a total 20 μL solution. The RT-qPCR reaction was then carried out according to the following protocol: RT at 45°C for 15 min, RT inactivation and GoTaq® activation at 95°C for 2 min, followed by 40 cycles of 15-second denaturation at 95°C, 60-second annealing/extension. The plate was read after each cycle.

The RT-qPCR data were analyzed using the CFX Manager Software (BioRad, Hercules, CA, USA). The cycle threshold (Ct) values were determined by the default algorithm in the CFX Manager Software. Each run contained positive and non-template controls. Extraction RNA blanks were included monthly for field and RNA. Calibration curves (0 to 5-log range) were generated with tenfold serial dilutions of SARS-CoV-2 positive control (IDT, Coralville, IA, USA) in the range from 200,000 to 2 genome copies (gc)/μL. Correlation coefficients (R2) > 0.99 were obtained for all calibration curves, with 90% to 100% amplification efficiencies. Each qPCR assay RNA elution had a limit of detection (LoD) of > 4 gc/μL, showing more than 50% positive signal, with Ct values of the lowest-dilution positive control.

For the endogenous biomarker target, pepper mild mottle virus (PMMoV) was used for concentration method validation (i.e., samples positive for PMMoV but negative for SARS-CoV-2 were considered under LoD for SARS-CoV-2 virus; if both were negative, we reprocessed the samples for confirmation). For the recovery control, human coronavirus-OC43 (HCoV-OC43) was used to evaluate the recovery rate in the wastewater due to its similar envelope structure. The SARS-CoV-2 recovery efficiency was carried out as described previously by Gharoon et al.^22^

### 1.3 Library Preparation and Sequencing

Sequencing libraries for genotyping SARS-CoV-2 virus in wastewater were prepared by one of two methods. Wastewater samples for variant analysis (all WRFs and sewershed) collected from November 2021 to July 2022 were processed as previously described by our group in Hartley et al.^23^ Briefly, RNA was linearly amplified into dsDNA, sheared, and ligated to Illumina-compatible sequencing adapters with the QIAGEN QIAseq FX Single Cell RNA Library Kit. These PCR amplicons were sequenced as 2×151. These libraries were then enriched for SARS-CoV-2 sequences with an Arbor Biosciences library enrichment kit and SARS-CoV-2 specific enrichment probes. These libraries were sequenced as 2×60. These libraries were sequenced as paired reads with the NextSeq 2000 P2 100-cycle sequencing kit. Due to decreasing coverages (described in *Limitations*) as the study period progressed, wastewater samples collected between August 2022 and November 2022 (and select samples acquired before August 2022) were processed with the QIAGEN QIAseq DIRECT SARS-CoV-2 kit according to manufacturer instructions and sequenced as paired reads with a NextSeq 2000 P1 300 cycle sequencing kit. Depending on kit availability, RVP samples were either sequenced as 2×151 or 2×60.

For the identification of respiratory pathogens in the wastewater, RNA was extracted from samples with Ct values (range 35–37), as previously described.^12^ Briefly, samples were treated with DNase I (QIAGEN, Inc., Germantown, MD, USA) for 30 minutes at room temperature before concentrating through RNeasy Minlute spin columns (QIAGEN, Inc., Germantown, MD, USA). Once concentrated, samples were converted into Illumina-compatible sequencing libraries using either the Respiratory Pathogen ID/AMR Enrichment Panel kit or the Respiratory Virus Oligo Panel (RVOP) according to the manufacturer’s protocols (Illumina, Inc.). Briefly, RNA was first denatured using the Elute, Prime, Fragment High Concentration Mix (EPH3) for 5 minutes at 65°C. Hexamer-primed RNA fragments were then reverse-transcribed to produce first-strand complementary DNA (cDNA). Second-strand synthesis was performed to complete the cDNA. AMPure XP beads were used to clean up the cDNA for tagmentation. Following the tagment step, premixed Index 1 (i7) and Index 2 (i5) adapters (Illumina, Inc.) were added to the sample and subjected to 16 PCR cycles to amplify the tagged cDNA and incorporate the adapters. Samples were again cleaned with AMPure XP beads. The cDNA was then normalized and consolidated into one-plex samples using undiluted libraries for overnight hybridization at 58°C. Enrichment of SARS-CoV-2 and RVPs was done via the Respiratory Pathogen ID/AMR Enrichment or RVOP Enrichment Oligos (labeled with biotin), captured with streptavidin-coated beads and washed. The resulting enrichment pools were quantified and normalized using High Sensitivity D1000 ScreenTape® and TapeStation Analysis Software 3.2. Sequencing was performed using an Illumina NextSeq mid-output (2×75) or NextSeq 2000 P2100 cycle (2×50). The generated FASTQs from the sequencing reaction were subjected to the detection of RVP signatures.

### 1.4 Bioinformatics and Data Analysis

Single-end (SE) or paired-end (PE) FASTQ files generated from the Illumina sequencing were analyzed using a custom bioinformatics pipeline publicly available on GitHub. Details about the pipeline and setup can be obtained from its GitHub page (https://github.com/Nevada-Bioinformatics-Center/snakemake_freyja_covidwastewater). This pipeline starts by trimming the reads using fastp (https://github.com/OpenGene/fastp) to remove poor-quality bases and adapter contamination. Then, the reads are classified using Kraken2 using its standard taxonomic database (https://github.com/DerrickWood/kraken2) to assess the quality and content of organisms sequenced in the wastewater sample. The trimmed reads are mapped to the Wuhan-Hu-1 reference (MN908947.3) using minimap2 (https://github.com/lh3/minimap2), and the resulting BAM files are assessed for quality control (QC) using Qualimap (http://qualimap.conesalab.org/). A combined QC report of all samples, including fastp trimming, Kraken2 classification, and Qualimap, is then generated by MultiQC (https://github.com/ewels/MultiQC). Freyja (https://github.com/andersen-lab/Freyja) runs on each sample to recover the relative lineage abundances from the mixed SARS-CoV-2 samples using the mapped BAM files. Freyja identifies the total coverage per sample and the abundances for each type of variant derived from the UshER global phylogenetic tree. Lastly, an aggregated report is generated, which includes viral concentration data and the relative abundances of each SARS-CoV-2 variant. This report is then used to visualize and compare the data across different sites over time and to assess the correlations between viral concentration, the presence of AMR genes, and the relative abundances of different SARS-CoV-2 variants.

### 1.5 Detection of RVPs and AMR Genes

We used an open-source IDseq pipeline (v3.7, https://czid.org/) to analyze the presence of RVPs and AMR genes.^24^ Briefly, the pipeline performs subtractive alignment of the human genome (NCBI GRC h38) using STAR (v2.5.3),^25^ followed by quality filtering with subsequent removal of cloning vectors and phiX phage using Bowtie2 (v2.3.4).^24^ The identities of the remaining microbial reads are then queried against the NCBI nucleotide (NT) database using GSNAP-L in the final steps of the IDseq pipeline.^24,26^ After background correction and filtering, retained taxonomic alignments in each sample were aggregated at the genus level and sorted by abundance, measured in nucleotide reads per million (NT-rPM), independently for each sample.

## RESULTS

In this study, we aimed to determine the presence and concentrations of SARS-CoV-2 in wastewater samples collected from the Reno-Sparks metropolitan area and understand how the concentration of SARS-CoV-2 in wastewater correlates with any emerging variants of the SARS-COV-2 virus. We also conducted metagenomic analyses to detect other respiratory viruses and genes associated with AMR in the samples. We collected 175 wastewater samples from three wastewater treatment plants (WRFs) and seven regional sites in the Reno-Sparks metropolitan area between November 2021 and November 2022. The recovery rate of the viral genomic RNA was about 23% from the wastewater matrix. The concentration of SARS-CoV-2 gene copies ranged between the 4 gc/μL (LoD) and 8.48 × 10^5^ gc/L (N1 gene) and 3.32 × 10^6^ gc/L (N2 gene). We detected viral genomes with Ct values as low as 38.54 (about 1.35 × 10^4^ gc/L).

### 1.6 SARS-CoV-2 Concentrations in Wastewater

Measuring the concentration of SARS-CoV-2 in wastewater can provide insights into the prevalence of COVID-19 in a community and help predict future outbreaks. By analyzing the concentration of SARS-CoV-2 in wastewater samples collected over a 12-month period, we were able to track changes in the virus concentration of COVID-19 in the area. Longitudinal analysis of genomic RNA copies of the SARS-CoV-2 nucleocapsid genes, N1 and N2, are represented for TMWRF (**Figure 1** [logarithmic gc]) and all other sites (**Supplementary Figure 1** [linear gc/L]). At TMWRF, there were relatively low concentrations of N1 (log 3.3 ± 0.20) and N2 (log 3.67 ± 0.27) in the wastewater from November 2021 until early December 2021 (Delta, pre-Omicron period) (mean ± standard deviation [SD]).

**Figure 1.**
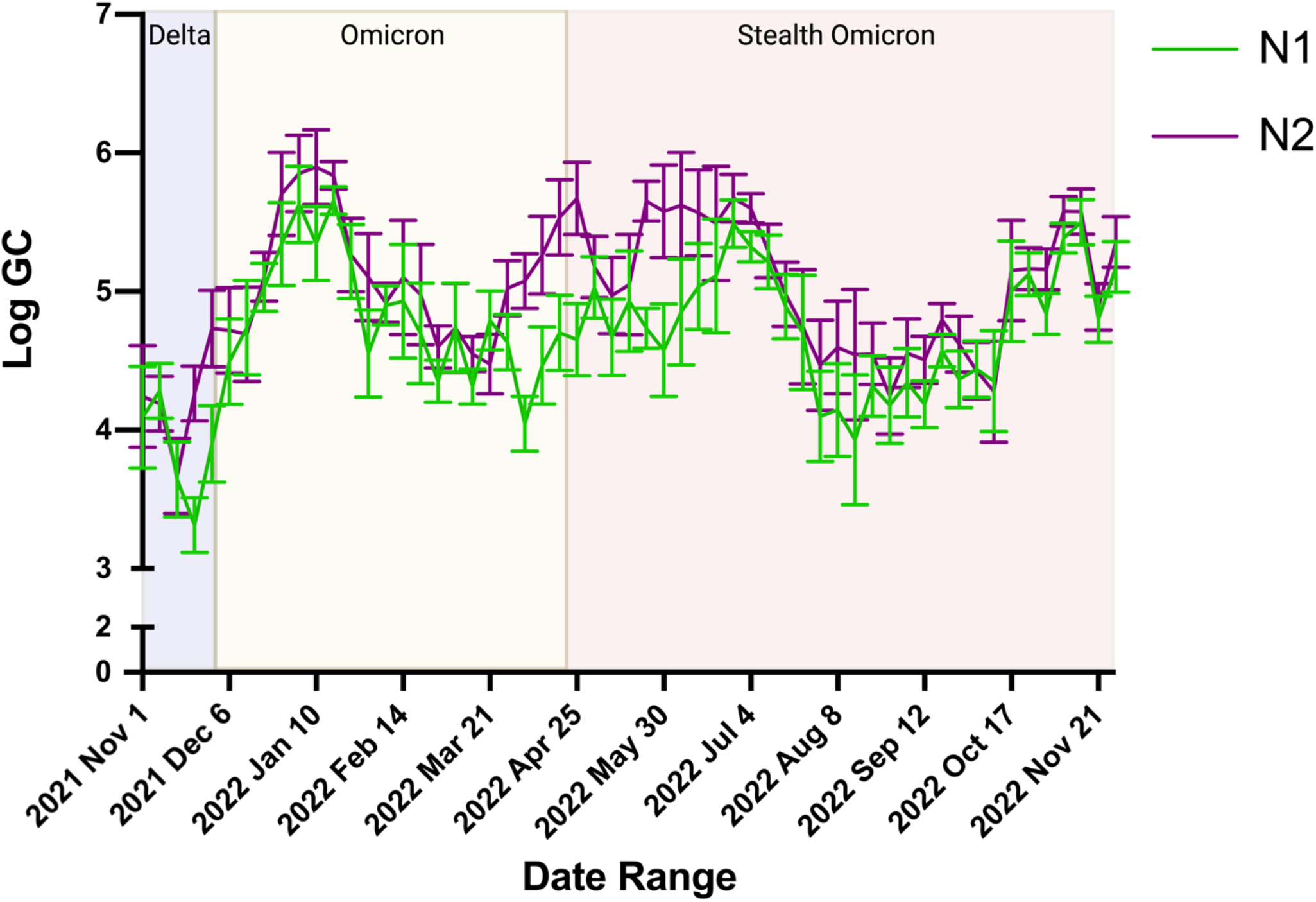
Longitudinal Analysis of Genomic RNA of SARS-CoV-2 Nucleocapsid Genes, N1 and N2. Influent was collected from November 2021-November 2022. RT-qPCR results for N1 (green) and N2 (purple) are plotted over time. Weekly mean logarithmic N1 and N2 GC is plotted with error bars representing standard deviation. Figure created with BioRender.com.

We found that the concentration of SARS-CoV-2 N1 and N2 in the wastewater varied over time. During the Omicron wave (December 2021-mid January 2022), the concentration rose sharply and then returned to early December levels. A similar pattern was observed during the Stealth Omicron wave (April-early August 2022). However, in the later months of the study (October-November 2022), we saw an increase in the N1 and N2 concentrations of SARS-CoV-2 in the wastewater. This pattern was also seen at other WRFs and sewershed. (**Supplemental Figures 1A-E**).

### 1.7 Detection of SARS-CoV-2 Variants in TMWRF Sewershed

We performed SARS-CoV-2 variant analysis at TMWRF using the updated Freyja variant profiling classification analysis to classify the variants (snakemake pipeline) (**Figure 2**). By depicting the variant data in a chronological manner, we visualized SARS-CoV-2 evolutionary inflection points (i.e., the presence of a variant [occurring for short – or long-term] preceding the generation of a new variant(s) that then proceed to become predominant). These inflection points were determined to be Dec 6^th^, 2021 (beginning of the Omicron wave) with B.1.1.529, and April 8^th^, 2022 (beginning of the Stealth Omicron wave), with BA.2.12.1. Thus, following these variant introductions, predominant variants during each respective wave were identified as described below. Further, these inflections can also be visualized where significant increases in N1 and N2 (log gc) proceed the Omicron and Stealth Omicron waves, respectively (**Figure 1**).

**Figure 2.**
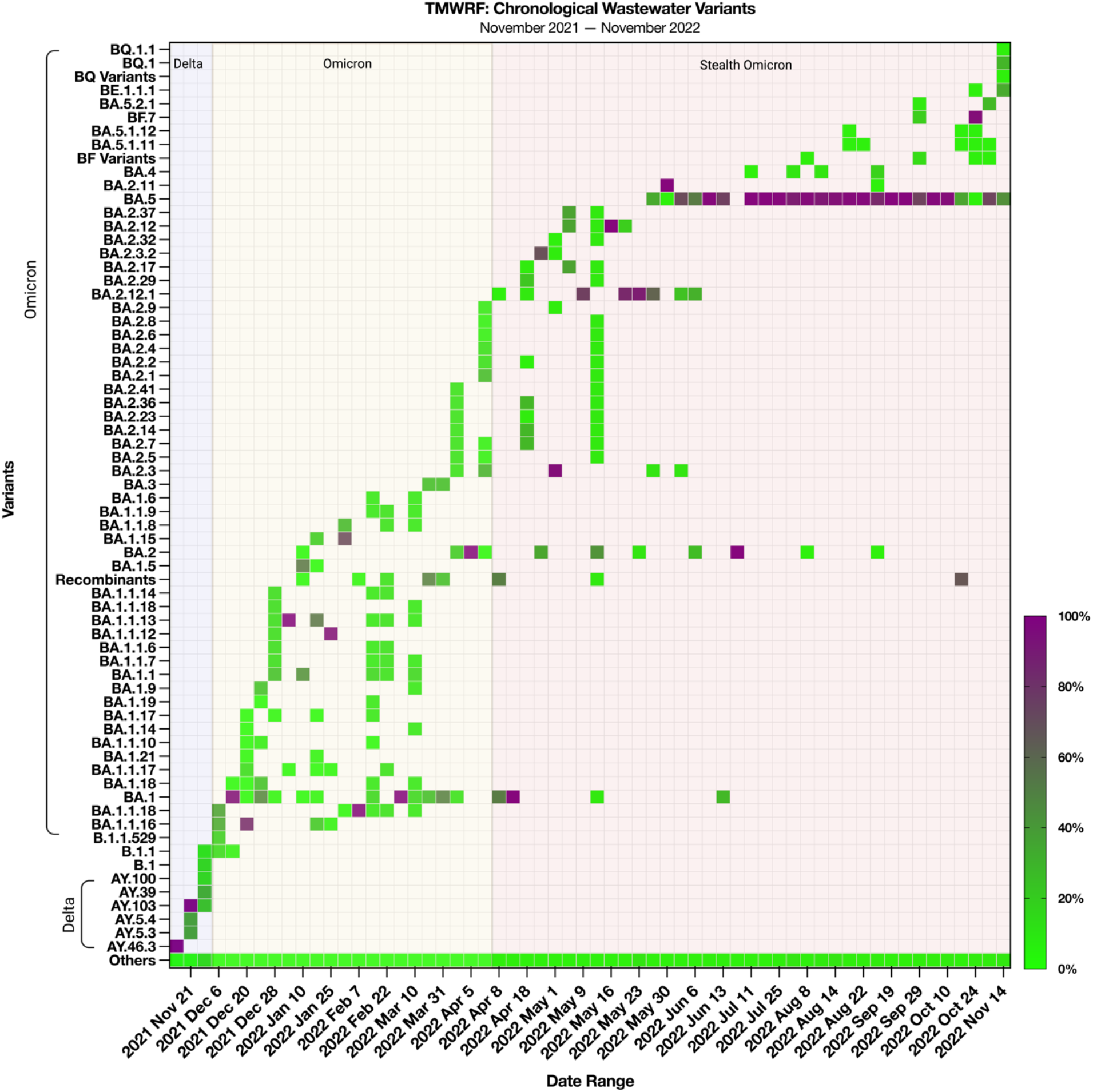
Chronological Detection of SARS-CoV-2 Variants in TMWRF Sewershed. Influent was collected from November 2021-November 2022. The relative proportion of variants was determined by Freyja, a SARS-CoV-2 variants analysis pipeline. B.1.1.529 (Omicron) was first detected on Dec. 6^th^, 2021, before the identification of a clinical case of Omicron in Washoe County. BA.2.12.1 (Omicron) was first detected on Apr. 8^th^, 2022. Figure created with BioRender.com. **\***TMWRF geographic location is represented in **Figure 4**.

During the Omicron wave (December 2021 through January 2022), the variants with great than 75% prevalence were BA.1, BA.1.1.16, BA.1.1.13, BA.1.1.12, BA.1.1.18, BA.2, and BA.2.3 (**Figure 2**). Other variants, including BA.3 and its subvariants, were detected during this time period but were not predominant (e.g., BA.3 was only found in a few weekly samples at a prevalence of less than 25%). The introduction of the BA.2.12.1 variant led to a new Stealth Omicron wave (April 2022 through October 2022). The predominant variants (greater than 75% prevalence) were BA during this wave.2.12.1, BA.2.3.2, BA.2.12, BA.2.11, BA.5, and BA.5.10. BA.5 subvariants were the most predominant during this time period, but their prevalence began to decrease in October 2022, as the prevalence of BQ.1 and BE.1.1.1 variants increased. (**Figure 2**). BA.4 and its subvariants were detected during the Stealth Omicron wave but were not predominant. As plotted, we show respective subvariants with at least two weeks of detection (i.e., variants detected once were reclassified to their parent BA.X variant group). In our most recent TMWRF influent samples, we detected BQ (BQ.1 [24.88%] and BQ.1.1 [0.66%]), BF (mainly BF.7 and subvariants [96.3%]), and BE (BE.1.1.1 [29.68%]) variants. BQ.1 and BE.1.1.1 have established a combined prevalence of 54.31%, surpassing the declining BA.5 variants (41.52%).

### 1.8 Detection of RVPs in TMWRF Sewershed

In addition to analyzing the SARS-CoV-2 variants, we also performed metagenomic analysis to identify other co-occurring human respiratory disease-causing pathogens which allowed us to assess the prevalence of multiple respiratory viruses in the community. (**Figure 3**). The results of the metagenomic analysis showed that the prevalence of SARS-CoV-2 was consistently high throughout the study period, while the prevalence of other viruses had a seasonal pattern. For example, the prevalence of human Mastadenovirus increased during the early winter months, while Paraechovirus and Parvovirus (NIH-CQV) had a higher prevalence during the summer months. Low levels of enteric viruses, such as Mastadenovirus (F) and Enterovirus sp. (B and C), were detected throughout the year as expected. Other human respiratory disease-causing viruses that were detected in the metagenomic analysis included Paraechovirus (A), Mastadenovirus sp. (A, B, C, D, E, and G), Influenza (A and C), Rhinovirus sp. (A, B, and C), human Polyomavirus (1), HMO Astrovirus (A), and human Bocavirus.

**Figure 3.**
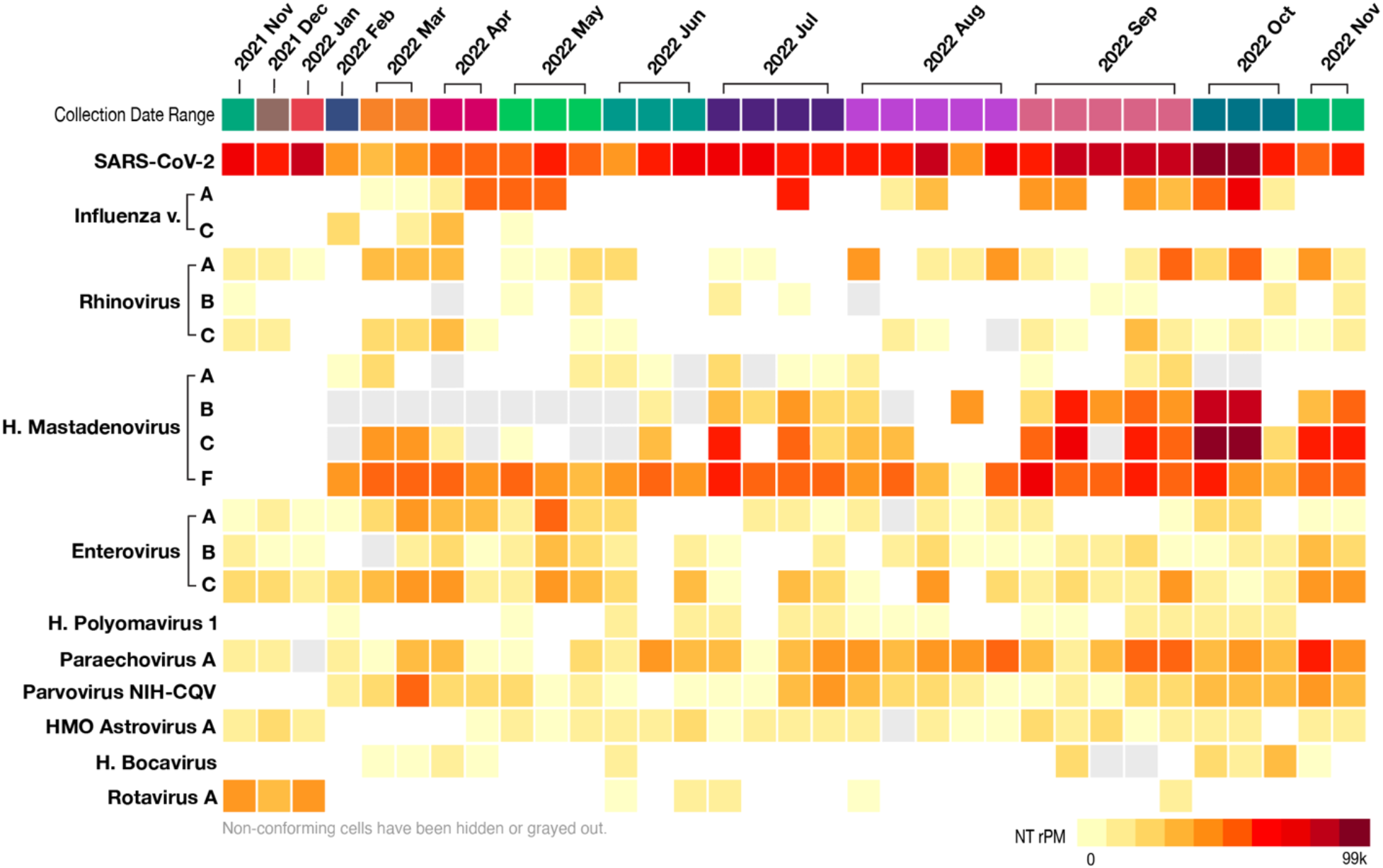
RVP Metagenomic Analysis of TMWRF Sewershed. Influent was collected from November 2021-November 2022. The relative proportion of SARS-CoV-2 and other RVPs were conducted through sequencing of genomic RNA using an RVP kit (Illumina). The sequences were analyzed through the Pathogen Detection tool of CZiD.org. A heatmap was generated based on the number of total reads per million (NT rPM) of each pathogen detected relative to their overall abundances.

### 1.9 Detection of AMR Genes in TMWRF Sewershed

To investigate the presence of AMR genes in the TMWRF region, we conducted a secondary analysis to identify genes that confer resistance to various antibiotic classes (**Table 1**). We detected AMR genes that confer resistance to macrolide, beta-lactamase, tetracycline, sulfonamide, glycopeptide, fluoroquinolone, trimethoprim, aminoglycoside, phenicol, and colistin antibiotics. These AMR genes had coverage ranging from 2.34% to 94.28%, and AMR gene reads (per million) ranging from 0.01 to 1911.99 RPM. The most predominant AMR genes in our samples belonged to the macrolide and tetracycline antibiotic classes, followed by beta-lactamase and aminoglycoside. The sulfonamide antibiotic class had two AMR genes detected at a prevalence of greater than 90%. The glycopeptide, fluoroquinolone, trimethoprim, phenicol, and colistin antibiotic classes had fewer AMR genes detected, with 1 or 2 dominant AMR genes in the respective antibiotic class (40-77% prevalence).

**Table 1.** Antimicrobial Resistance Gene Analysis from Wastewater Influent.

| **Antibiotic Class** | **Gene** | **Sample Counts** | **Avg. Coverage of AMR Gene (%)** | **Avg. RPM of AMR Gene** |
| --- | --- | --- | --- | --- |
| **Macrolide** | **MphE** | 31 | **89.43** | 530.71 |
|  | **MsrE** | 31 | **88.66** | 1193.98 |
|  | **MefA/Mel** | 31 | **66.34** | 105.50 |
|  | MsrD | 31 | 53.02 | 0.94 |
|  | **ErmF** | 30 | **83.78** | 382.78 |
|  | **ErmG** | 30 | **79.10** | 92.04 |
|  | Mef(B) | 29 | 38.23 | 0.52 |
|  | **ErmB** | 28 | **70.91** | 163.25 |
|  | **LnuC** | 28 | **58.07** | 71.07 |
| **Beta-lactamase** | **OXA-7** | 31 | **73.61** | 94.27 |
|  | **OXA-2** | 31 | **62.42** | 204.32 |
|  | **TEM-1D** | 31 | **67.07** | 104.44 |
|  | **CfxA** | 30 | **70.58** | 1.11 |
|  | OXA-211 | 27 | 49.24 | 99.91 |
|  | OXA-5 | 22 | 27.93 | 1.86 |
|  | CARB-5/BlaRTG-2 | 19 | 33.71 | 12.59 |
|  | CEPH | 15 | 23.85 | 1.77 |
| **Tetracycline** | TetC | 31 | 36.30 | 0.52 |
|  | **TetQ** | 30 | **94.28** | 1483.71 |
|  | **TetX** | 30 | **86.70** | 974.86 |
|  | **TetW** | 30 | **81.13** | 1911.99 |
|  | **TetO** | 30 | **76.17** | 566.78 |
|  | TetM | 30 | 38.20 | 65.54 |
|  | TetB-P | 30 | 40.45 | 0.83 |
|  | TetA-P | 30 | 38.90 | 0.60 |
|  | Tet-32 | 29 | 38.42 | 126.82 |
|  | Tet-39 | 29 | 49.00 | 0.61 |
|  | **Tet-44** | 29 | **53.15** | 1.16 |
|  | Tet-40 | 26 | 27.79 | 1.62 |
| **Sulfonamide** | **Sull** | 30 | **64.05** | 173.47 |
|  | **SulII** | 28 | **52.85** | 77.65 |
| **Glycopeptide** | VanA-G | 16 | 20.97 | 0.08 |
|  | VanRD | 1 | 13.59 | 0.14 |
|  | VanTC | 2 | 3.33 | 0.05 |
|  | VanSA | 1 | 2.25 | 0.10 |
|  | VanYD | 1 | 8.15 | 0.05 |
|  | VanRF | 1 | 6.03 | 0.02 |
| **Fluroquinolone** | QnrVC5 | 13 | 48.67 | 21.10 |
|  | OqxB | 24 | 6.14 | 0.23 |
|  | QnrVC1 | 8 | 20.09 | 5.52 |
|  | QnrD | 8 | 38.02 | 22.66 |
| **Trimethoprim** | DfrA5 | 13 | 46.30 | 21.41 |
|  | DfrA16 | 8 | 32.75 | 3.65 |
|  | DfrA3 | 6 | 23.79 | 0.81 |
|  | DfrA1/Dfr1 | 6 | 39.30 | 6.84 |
| **Aminoglycoside** | Aph3-III | 30 | 49.13 | 116.08 |
|  | **ANT(6)-Ib** | 27 | **54.03** | 67.68 |
|  | ANT(6)-Ia | 24 | 42.64 | 43.85 |
|  | Aac3-I | 20 | 33.91 | 17.87 |
|  | APH(3")-Ia/AphD2/AphE | 15 | 20.76 | 0.14 |
| **Phenicol** | FloR | 3 | 5.79 | 0.06 |
|  | Cmr | 2 | 9.86 | 0.03 |
|  | CmlB1 | 2 | 2.77 | 0.04 |
|  | CatA2 | 1 | 17.13 | 0.03 |
|  | CatB | 1 | 4.09 | 0.11 |
| **Colistin** | MCR-4.1/MCR-4 | 2 | 3.88 | 0.03 |
|  | MCR-1.1/MCR-1 | 1 | 2.34 | 0.01 |

\*AMR genes >50% Avg. Coverage Bolded

### 1.10 Detection of SARS-CoV-2 Variants in RSWRF, STMWRF, and Sub-Sewersheds

We analyzed the prevalence of SARS-CoV-2 variants at all the tested sites to understand the distribution and evolution of the virus in the community. The results showed that all the sites had a similar prevalence of variants, with slight variations in the order of appearance (**Figure 4**). BA.5 was the predominant variant at TMWRF from April 8^th^, 2022 (inflection point for the Stealth Omicron wave). However, our sample collection at the non-TMWRF sewershed began on May 3^rd^, 2022. RSWRF showed an increasing prevalence of BA.5 (range 43-99%) over the collection period, with a single sample with a high prevalence of BA.2.12.1 (33.64%) on July 13^th^, 2022. The first overall detection of BF and respective subvariants occurred on August 3^rd^, 2022, at RSWRF (4.32%), with subsequent detections at STMWRF on October 3^rd^, 10^th^, 24^th^, and November 7^th^ (0.35%, 0.44%, 0.44%, 65.81%, respectively), Travel-Influenced sites on August 9^th^ and 10^th^, 2022 (10.76%, 55.45%) and Elementary Schools on October 19^th^, 2022 (74%). There were no detections of BF variants at Sub-Neighborhoods. BG and respective subvariants were detected in the effluent collected from the Elementary Schools on May 3^rd^, 2022 (97.4%), followed by Sub-Neighborhoods on Jun 14^th^, 2022 (0.16%), and at STMWRF on Jun 20^th^, 2022 (0.65%). There were no detections of the BG variants at TMWRF. There was only one low trace detection of BE and respective subvariants in STMWRF on August 29^th^, 2022; however, there has since been no detection of BE at any non-TMWRF site (as of October 24^th^, 2022). BQ and respective subvariants were detected as early as October 31^st^, 2022 in STMWRF, confirming the introduction of this new variant.

**Figure 4.**
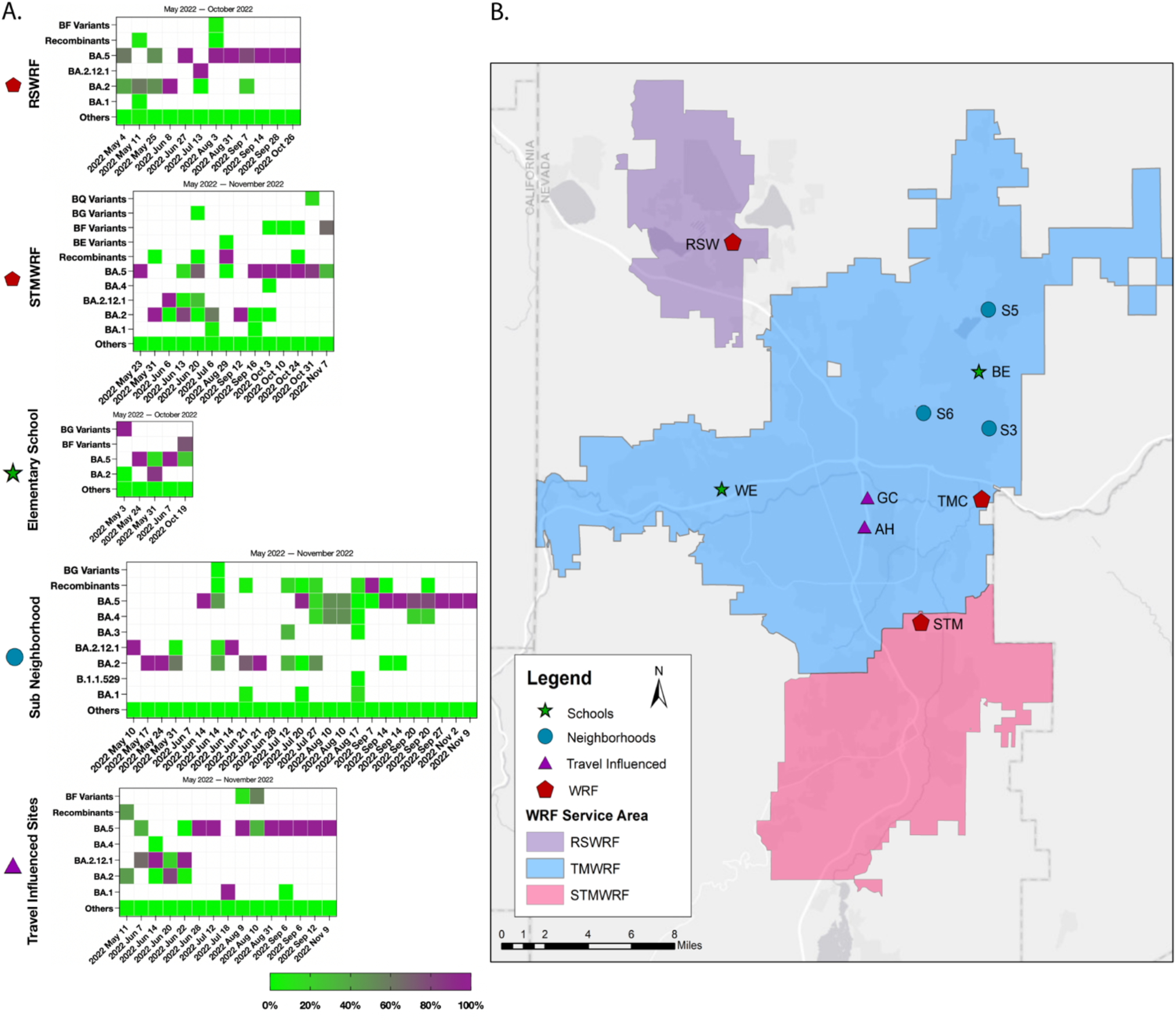
Detection of SARS-CoV-2 Variants in STMWRF, RSWRF, and Other Sub-sewershed. Influent was collected from May 2022 – October 2022 at two WRFs and 3 sewershed (A). A map of the Reno-Sparks metropolitan area depicting WRFs and sewer shed locations and servicing areas is shown (B). Collective occurrences of the main variants of SARS-CoV-2 Delta and Omicron (e.g., AY.X, BA.1.X, BA.2.X, BA.3.X, BA.4.X, BA.5.X, BE.X, BF.X, BG.X, and BQ.X) were calculated. The relative proportion of variants was determined using Freyja, SARS-CoV-2 variants analysis pipeline. BA.2.12.1 was excluded from summation to depict the significance of detection. *TMWRF variant data is shown in **Figure 2**

## DISCUSSION

WBE for SARS-CoV-2 analysis is a recent global endeavor to mitigate low testing frequencies and reporting information to gather more comprehensive data on the current status of COVID-19 in a community.^20,27-30^ Moreover, WBE has been effective in detecting key variants responsible for the evolution of SARS-CoV-2 infectivity and pathogenicity.^13,15,20,31^ In this study, we analyzed multiple WRFs and local collection sites in the Reno-Sparks metropolitan area using RT-qPCR to quantify the presence of SARS-CoV-2 and its variants. To contribute additional insight to the literature, the present study sought to independently assess different regional WRFs and local collection sites for the quantification of SARS-CoV-2 and respective variants via frequent sample collection and by using an up-to-date variant classification bioinformatics analysis. Furthermore, our study is strengthened through secondary analysis and identification of other respiratory disease-causing viruses and AMR genes not included in other respective studies. Our findings provide valuable information on the COVID-19 status in the Reno-Sparks area and demonstrate the usefulness of WBE in detecting circulating viruses. Our data showed detections of new variants. We noted a recent rise in viral genome concentrations that was not reflected in the number of clinically diagnosed cases, possibly indicating a discrepancy between testing and reporting at the community level.

Our RT-qPCR data on the N1 and N2 genes in wastewater show early signs of all new variants (**Figure 1**), and continual monitoring for variants allowed for early and specific detection of new VOIs and VOCs. WBE is particularly useful in times of low testing frequencies and can provide comprehensive, up-to-date data on the COVID-19 status of a community, including individuals who are not being tested. This is particularly valuable in rural areas where access to testing may be limited. Previous research has shown that SARS-CoV-2 cases tend to spike in the summer and winter months, which may lead to the emergence of new variants, which, based on the current pattern, could be driving the BE, BF, or BQ variants.^32,33^ We also note a recent rise in viral genome concentrations; however, we do not see the same trend in clinically diagnosed cases, alluding to a discourse between testing and reporting at a community level. Thus, WBE of SARS-CoV-2 is once again proving to be an effective tool during times of low testing frequencies and reporting, and further, can be used as a more effective and inclusive (i.e., acquisition of virus prevalence amongst testing and non-testing persons) diagnostic in rural settings to provide public health and city officials of up to date community data, especially as the process becomes increasingly more streamlined and broadly accepted.^34,35^

### 1.11 Variants and Genomic Surveillance

There are nationally recognized entities that exist to provide updated information on variants of interest (VOIs), variants of concern (VOCs), and variants being monitored (VBMs) (e.g., Centers for Disease Control [CDC], World Health Organization [WHO]).^36,37^ Current CDC guidelines report pre-Omicron variants as VBMs (e.g., Alpha, Delta, Wuhan strains). One variant is currently reported as a VOC (B.1.1.529); however, no VOIs are currently established.^37^ During the beginning of this study (November 2021), almost all the signatures detected were of the Delta (AY) variant, which got replaced following the introduction of the B.1.1.529 Omicron variant (**Figure 1**). Since their disappearance in the wastewater, we have not re-detected their signatures (**Figure 2**). This was a similar WBE result seen by Galani et al. in Greece (e.g., the Delta variant being replaced by B.1.1.519, signifying the start of the Omicron wave).^38^ The B.1.1.529 (Omicron) variant was first detected in our study on December 6^th^, 2021, which preceded the Omicron wave by one week, and consisted of predominantly BA.1, BA.1.1.18, BA.1.1.12, BA.1.1.13, and BA.2 variants.

Preceding the Stealth Omicron wave of BA.2 and respective subvariants, we saw the highest prevalence of BA.2.12.1, which we then hypothesized to be the predominant variant in all our study sites to precede BA.5 in high concentrations (**Figure 2**). This followed early data reporting that US sequences submitted to GISAID accounted for 26% of BA.2.12.1 and that BA.4 and BA5 comprised more than 90% of the genomes sequenced in South Africa.^39^ Furthermore, BA.2.12.1 has been observed to overcome immunity from earlier Omicron infections, suggesting that it may cause re-infections and increase infectivity.^40,41^ In our TMWRF samples, which represent the most extensive collection area, BA.2.12.1 was detected seven weeks prior to the first sample containing BA.5. Although sample collection dates at non-TMWRF sites are limited, we did see BA.2.12.1 as the predominant variant at two other sites (Sub-Neighborhood and Travel-Influenced Sites) preceding BA.5; however, BA.2.12.1 was predominant alongside BA.5 in two other sites (STMWRF, RSWRF) (**Figure 4**). In contrast to B.1.1.529, which had a brief prevalence preceding the BA.2 Omicron wave, BA.2.12.1 had a longer time course before BA.5.

The detection of BF and respective subvariants in our sub-sewershed sites on August 3^rd^, 2022 is noteworthy (**Figure 4 and Supplemental Figure 2**). Despite the early detection and rapid drop of these BF and respective subvariants, these variants have the potential to become the predominant strain in the region, as evidenced by the increasing prevalence observed in our largest site (TMWRF): August 8^th^, 2022, with BF.8 (0.8%), followed by BF.7 (15%) on September 29^th^, and BF.7 (96.35%) on October 24^th^, 2022. The last TMWRF sample with the BF variant was on October 31^st^, 2022, with BF.4 (0.44%). Other sub-sewershed sites also showed detections of BF variants, with RSWRF detecting BF.3.1 (4.03%) and BF.1 (0.21%) on August 3^rd^, 2022, and Travel-Influenced sites detecting BF (10.76%) and BF.2, BF.4, and BF.18 (26.14% each, respectively) on August 9^th^, 2022. Elementary schools also detected BF.12, BF.7, and BF.28 (24.5% each, respectively) on October 19^th^, 2022, where these BF subvariants were more predominant than BA.5. These data collectively indicate that BF may also become the predominant variant in the region, especially when considering rising abundance at TMWRF.

Our TMWRF data suggests that BQ.1 and BE.1.1.1 are beginning to establish strong predominance in the region, with the exception of STMWRF, which continued to show a higher prevalence of BA.5.2 and BA.5.2.1 subvariants in recent samples (October 24^th^ – November 7^th^, 2022). The higher prevalence of BQ.1 is in accordance with the CDC COVID-19 variant foresight and prevalence tracker, which uses a NOWCAST model to estimate and predict proportions of circulating variants based on recently circulating variant proportions, is predicting that BQ.1, BQ.1.1, and BF.7 variants to become the predominant future variants in the region including NV (Region 9).^42^

### 1.12 RVP and AMR Analysis

This WBE analysis consistently detected high levels of SARS-CoV-2 throughout the year compared to other respiratory viruses (e.g., Influenza), which varied seasonally. This may suggest that SARS-CoV-2 was constantly present in the community but caused fewer clinical cases in the summer months. Cold temperatures are known risk factors for respiratory infections, particularly in the winter, when they can cause irritation and inflammation of the airways, making it easier for respiratory viruses to enter and infect the body (e.g., irritation and inflammation of the airways, making it easier for respiratory viruses to enter and infect the body).^43,44^ Furthermore, studies have found that respiratory infections are higher during the winter months as people are more likely to be exposed to cold temperatures (e.g., the rate of Influenza infections in the US was significantly higher at colder temperatures and climates compared to warmer temperatures and climates).^45,46^ Another study associated lower temperatures with higher rates of SARS-CoV-2 infection.^32^

One possible reason for lower temperatures associating with the increased spread of respiratory viruses is the disruption of extracellular vesicle swarms.^44^ These swarms contain contents produced by cells in response to the stress of stimuli (e.g., cold temperatures). They carry viral particles and other pathogens, which are released through exhalation and coughing.^43^ A recent study found that cold exposure may impair extracellular vesicle swarm-mediated nasal antiviral immunity, a physiological process involving innate Toll-like receptor 3 (TLR3) dependent antiviral immunity.^44^ WBE, paired with secondary analyses for respiratory disease-causing viruses, provides a comprehensive assessment of the total microbial burden in a community and can inform healthcare responses.^47,48^

The introduction of antibiotics has played a crucial role in the treatment of diseases globally, but the simultaneous delivery of these antibiotics has allowed microbes to develop antibiotic resistance, reducing their therapeutic effect in humans and leading to difficulties in the management of many infectious diseases.^49^ In addition, the rise in AMR (and associated AMR genes) is responsible for over 700,000 worldwide deaths yearly.^50^ Brumfield et al. performed secondary metagenomic, metatranscriptomic, and targeted SARS-CoV-2 analyses on wastewater samples as cases were rising, and found the presence of other pathogens and important spike mutations, as well as potential coinfection insights and AMR genes.^7^ Similarly, our study identified significant macrolide and beta-lactamase AMR genes.

A recent study investigated the levels of beta-lactam antibiotics via the mecA gene by comparing wastewater samples from hospitals and other healthcare facilities to other sources, and a higher prevalence of the MecA gene in hospitals and healthcare facilities.^51^ In our study, the most abundant beta-lactamase AMR genes were MphE and MsrE (89.43% and 88.66%, respectively), suggesting that the increased use of antibiotics may be related to secondary bacterial infections associated with SARS-CoV-2 and may have implications for changes in the human gut microbiome.^52,53^ While we found high levels of AMRs for many classes of antibiotics we also identified classes of antibiotics with lower levels of detected resistance, which may have a potential therapeutic effect against antibiotic-resistant pathogens.

### 1.13 Study Limitations

WBE provides valuable insight into regional virus prevalence and can provide data into circulating viral variants and antimicrobial resistance genes. However, it does not provide specific information on persons infected, and tracing efforts are difficult as wastewater samples are pooled at WRF sites (unless a specific sub-sewershed is used). WBE is a relatively new research approach to the public health and medical field, and a few ethical concerns must be considered. For example, there is the potential for discrimination or stigmatization of individuals or communities based on the results of the research, which may lead to social and economic consequences leading to undermined trust in research and public health efforts (e.g., if there is a high abundance of a particular RVP in a community, they may be targeted and blamed). Thus, anonymizing or aggregating data is important, as well as developing and following strict protocols for handling and sharing sensitive information. Furthermore, although the more significant sample regional area site (TMWRF) provides more generalizable data, there were slight differences between the smaller sub-sewershed, which indicates a decrease in the degree of generalizability provided by more extensive sample-area data. We also noted a gradual worsening in coverages in our variant analysis as the study period progressed, which required us to switch our amplification method (Qiagen). This could be partly due to the original enrichment oligos not working well due to evolution in the viral sequence, quality issues with the wastewater samples themselves, or degradation of reagents. Lastly, our sample collection, processing, and sequencing methodology were continuously analyzed and updated throughout the study period, which may contribute to some sample variation between early and recent data.

## CONCLUSION

### 1.14 Utility of Wastewater-Acquired Variant Data and Site-Specific Monitoring Strategies

Overall, our findings highlight the values of WBE for monitoring the presence and dynamics of SARS-CoV-2 in an urban setting. By collecting samples from multiple sites, we were able to identify regional differences in virus prevalence and detected the emergence of new variants which preceded the inflections of the two dominant Omicron waves and showed that near-proximity regional differences in virus prevalence exist (i.e., different variants are more prevalent in certain areas). The CDC-NOWCAST model’s prediction of the BQ.1, BQ.1.1, and BF.7 variants becoming the predominant future variants in the region aligns with our findings, further supporting the effectiveness of WBE in detecting and tracking the spread of SARS-CoV-2 and its variants. Moreover, this method does not rely on the requirement for persons to self-report or depend on testing infrastructure, as our samples are community-pooled wastewater, which provides better insight into community virus prevalence and supports WBE as a sufficient and acceptable alternative to clinical-based testing surveillance.

Our analysis also identified the presence of other RVPs and AMR genes, providing insight into the dynamics of SARS-CoV-2 co-occurring viruses, which can provide valuable information for healthcare teams. We observed a wave-like pattern of occurrence for respiratory disease-causing viruses aside from SARS-CoV-2 and Mastadenovirus F, as well as the identification of dominant AMR genes, such as MphE and MsrE. Additionally, our analysis revealed the seasonality of certain viruses, with SARS-CoV-2 and Influenza virus levels remaining consistently high throughout the year, while other viruses showed higher prevalence in specific seasons.

## Data Availability

All data produced in the present study are available upon reasonable request to the authors.

## AUTHOR CONTRIBUTIONS

MK, Conceptualization, Validation, Formal analysis, Investigation, Data Curation, Writing - Original Draft, Writing - Review & Editing; LL, Data Curation, Validation, Formal analysis, Investigation, Writing - Original Draft, Writing - Review & Editing; LH, Data Curation, Writing - Review & Editing; SHP, Data Curation, Validation, Investigation, Writing - Original Draft; KA, Data Curation, Validation, Investigation; MC, Data Curation, Validation, Investigation, Writing – Review & Editing;; TU, Writing - Review & Editing, Validation, Supervision; PDH, Data Curation, Validation, Formal analysis; HVG, Data Curation, Validation, Formal analysis, Writing - Original Draft, Writing - Review & Editing; JP, Data Curation, Validation, Formal analysis, Writing - Original Draft, Writing - Review & Editing; SCV, Project administration, Funding acquisition, Resources, Conceptualization, Validation, Formal analysis, Investigation, Data Curation, Writing - Original Draft, Writing - Review & Editing; KP, Project administration, Funding acquisition, Resources, Conceptualization, Validation, Writing - Review & Editing

## SUPPLEMENTAL INFORMATION

**Supplemental Figure 1.**
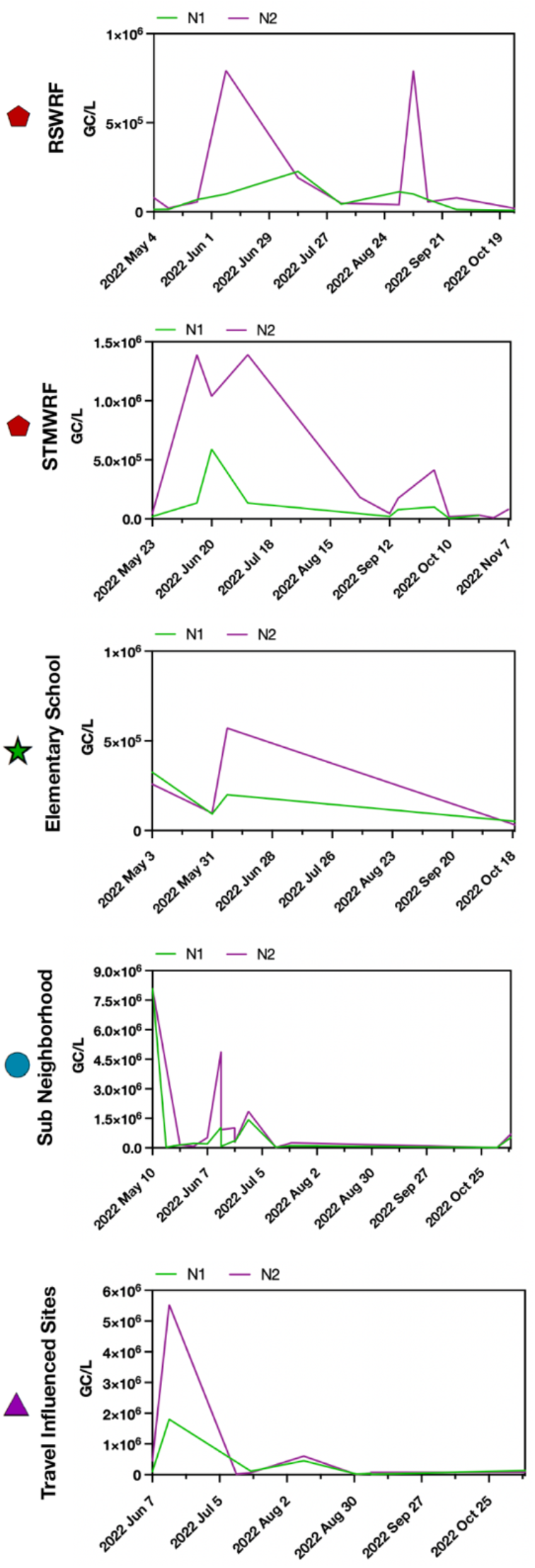
Longitudinal Analysis of Genomic RNA of SARS-CoV-2 Nucleocapsid Genes, N1 and N2, at Sub-Sewershed. Influent was collected from May 2022 - November 2022. RT-qPCR results for N1 (green) and N2 (purple) are plotted over time, subdivided by respective sewershed or WRF collection sites.

**Supplemental Figure 2.**
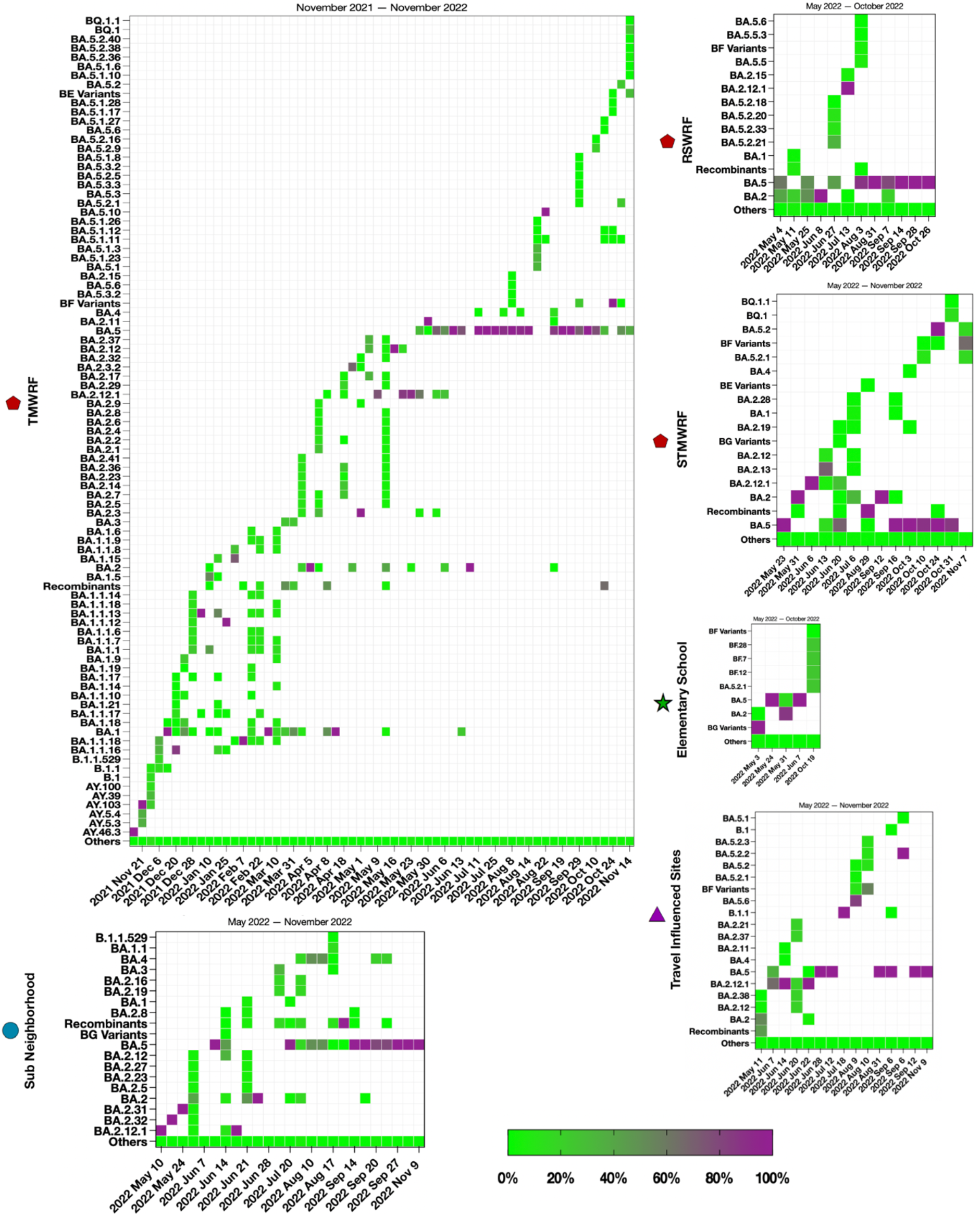
Expanded Detection of SARS-CoV-2 Variants in WRFs and Sub-Sewershed. Influent was collected from November 2022 – November 2022. Collective occurrences of SARS-CoV-2 Delta and Omicron (e.g., AY.X, BA.1.X, BA.2.X, BA.3.X, BA.4.X, BA.5.X, BE.X, BF.X, BG.X, and BQ.X) were calculated. The relative proportion of variants was determined using Freyja, SARS-CoV-2 variants analysis pipeline.

